# Risk factors, temporal dependence, and seasonality of human ESBL-producing *E. coli* and *K. pneumoniae* colonisation in Malawi: a longitudinal model-based approach

**DOI:** 10.1101/2022.08.11.22278326

**Authors:** Melodie Sammarro, Barry Rowlingson, Derek Cocker, Kondwani Chidziwisano, Shevin T. Jacob, Henry Kajumbula, Lawrence Mugisha, David Musoke, Rebecca Lester, Tracy Morse, Nicholas Feasey, Chris Jewell

**Affiliations:** Department of Clinical Sciences, Liverpool School of Tropical Medicine, Liverpool, United Kingdom; Centre for Health Informatics, Computing, and Statistics, Lancaster University, Lancaster, United Kingdom; Malawi-Liverpool-Wellcome Trust Clinical Research Programme, Kamuzu University of Health Sciences, Blantyre, Malawi; Centre for Water, Sanitation, Health and Appropriate Technology, Development (WASHTED), Malawi University of Business and Applied Sciences (MUBAS), Blantyre, Malawi; Department of Civil and Environmental Engineering, University of Strathclyde, Glasgow, United Kingdom; Global Health Security Department, Infectious Disease Institute, Makerere University, Kampala, Uganda; Department of Medical Microbiology, College of Health Sciences, Makerere University, Kampala, Uganda; College of Veterinary Medicine, Animal Resources and Biosecurity (COVAB), Makerere University, Kampala, Uganda; Conservation & Ecosystem Health Alliance, Kampala, Uganda; Department of Disease Control and Environmental Health, College of Health Sciences, Makerere University, Kampala, Uganda

## Abstract

**Background:** Antimicrobial resistance (AMR) represents an important threat to achieving the sustainable development goals in Sub-Saharan Africa (sSA). sSA is reported to have the highest estimated death rate attributable to AMR, with Extended-Spectrum Beta-Lactamase-producing Enterobacterales, such as *Klebsiella pneumoniae* and *Escherichia coli*, representing the greatest challenge. However, the dynamics of human colonisation with such bacteria in the sSA community setting are not well known. Inadequate water, sanitation and hygiene (WASH) infrastructure and associated behaviours are thought to play an important role in transmission of AMR-bacteria, and an improved understanding of the temporal dynamics of within-household transmission could help inform the design of public health policies that interrupt transmission of AMR-bacteria.

**Methods and Findings:** In this 18-month study, individuals from households in diverse areas of Southern Malawi were recruited and human stool samples were longitudinally collected. Using microbiological data and household surveys, we built a multivariable hierarchical harmonic logistic regression model to identify risk factors for colonisation with ESBL-producing *E. coli* and *K. pneumoniae*, reflecting household structure and temporal correlation of colonisation status between timepoints.

Important risk factors were identified, with men having a lower risk of becoming colonised with ESBL-producing *E. coli* (OR 0.786 CrI[0.678-0.910]) and the use of a tube well or a borehole as a water drinking source highly increasing the risk of becoming colonised (OR 1.550 CrI[1.003-2.394]). Coming into contact with standing water also appeared to be negatively associated with colonisation status (OR 0.749 CrI[0.574-0.978]). For ESBL-producing *K. pneumoniae*, having recently taken a course of antibiotics increased the risk of being colonised (OR 1.281 CrI[1.049-1.565]). We also found a negative association between eating from shared plates and colonisation with ESBL-producing *K. pneumoniae* (OR 0.672 CrI[0.460-0.980]). Finally, we detected a temporal correlation range of eight to eleven weeks, providing evidence that within-household transmission occurs within this time frame.

**Conclusions:** We suggest that interventions aimed at preventing transmission might have the best impact when targeted at the household-level and focused on a combination of improving WASH infrastructure and modifying associated behaviours. Additionally, we showed that antibiotic use is important when looking at colonisation with ESBL-producing *K. pneumoniae* and therefore infection prevention and control measures and antibiotic use and stewardship training could help in reducing transmission.

## Introduction

In 2015, the World Health Organisation (WHO) declared antimicrobial resistance (AMR) as one of the 10 global priority public health threats [1]. The rise of AMR internationally is endangering the achievement of the sustainable development goals, particularly those relating to health, poverty, food security and economic growth [2]. The latest estimates of the global burden of AMR showed that 4.95 million deaths were associated with bacterial AMR in 2019, among which 1.27 million deaths were attributable to bacterial AMR [3]. The death rate attributable to AMR was found to be highest in sub-Saharan Africa (sSA), where the leading pathogens were *Klebsiella pneumoniae, Streptococcus pneumoniae* and *Escherichia coli*, however large gaps in data availability were noted in sub-Saharan Africa [3]. Of particular note has been the rapid emergence of Extended-Spectrum beta-Lactamases (ESBL) in gram negative bacteria [4], potentially as a result of increased use of 3^rd^ generation cephalosporin antimicrobials in empirical treatment of bacterial infection in hospitals. Whilst studies have shown that the prevalence of infections caused by ESBL-producing Enterobacterales in sSA is high [5,6,7], little is known about asymptomatic colonisation with ESBL-producing Enterobacterales, a key step prior to infection in vulnerable patient groups. Learning more about asymptomatic colonisation in the community is therefore crucial in order to prevent transmission, and as a consequence, reduce drug-resistant infections.

Prior to 2016, no studies described risk factors for colonisation with ESBL-producing Enterobacterales among healthy individuals in the community in sSA[5]. More recently, recent antibiotic use (in the last weeks to months) was found to be a risk factor in a few community-based studies [8,9,10]. Other risk factors such as older age and previous hospital admission were also identified [8]. A further study found that higher income was associated with a higher prevalence of ESBL colonisation [11]. However, most of these studies focused on a specific subset of the community and not the general population. We cannot, therefore, be confident that the risk factors detected in these specific populations would be the same throughout the general population. This highlights the need for a community-based study within the general population to identify risk factors for human gut mucosal colonisation with ESBL-producing Enterobacterales. Moreover, although it is strongly believed that inadequacies in water, sanitation and hygiene (WASH) infrastructure and associated behaviours play an important role in transmission of AMR-bacteria [12], risk factors related to WASH in this context are still not well described. For example, only one study found that having private indoor access to drinking water was positively associated with ESBL colonisation [13].

To be able to reduce transmission of, and colonisation by, AMR-enteric bacteria in East and Southern Africa, we need to explore the dynamics of colonisation in order to tailor appropriate interventions; for example the role of seasonality in colonisation, and how long colonisation lasts after initial acquisition. A first step in understanding the temporal dynamics of within-household transmission is to determine how long a specific household is at risk of colonisation once one member has been colonised, this will help inform the design of public health policies that interrupt transmission of AMR-bacteria. Such interventions will be impactful at interrupting transmission of enteric pathogens more broadly.

Here, we describe an analysis of an 18-month longitudinal cohort study using microbiological, household and WASH surveys, where WASH refers to water, sanitation (containment of both human and animal faeces), hygiene and food hygiene. We fit a serially-correlated generalised linear mixed model, exploring household, individual and WASH risk factors for ESBL colonisation in settings with different degrees of urbanisation in Malawi.

This allows us to separate the effects of individual- and household-level risk factors from temporal effects we observe in our data.

## Methods

The Drivers of antimicrobial Resistance in Uganda and Malawi (DRUM) consortium was an interdisciplinary consortium working across urban, peri-urban and rural communities in Uganda and Malawi (www.drumconsortium.org). Its aim was to study AMR transmission in a One Health setting, sampling areas with different human and animal population densities, and different levels of affluence and infrastructure. DRUM was a repeated-measures study in which individuals, clustered into households, were sampled at four timepoints over 6 months. The detailed protocol is available at [14].

This analysis focuses on a subset of the data from the DRUM study, focusing on the 3 Malawi study areas: Ndirande, Chileka, and Chikwawa representative of urban, peri-urban, and rural demographics respectively. We model the presence or absence of gut-mucosal colonisation with ESBL *E. coli* and *K. pneumoniae* in individuals, and aim to detect associations with individual-level demographic and health characteristics, household-level WASH indicators, and the social context represented by the study area. In order to capture seasonality and household structure, we used a hierarchical multivariable harmonic regression, with temporal correlation at the household level. As described in the following sections, we first screened our candidate covariates through expert opinion and univariable logistic regression models, before taking these variables forward into our full modelling framework.

### Covariate selection

In order to investigate WASH practices at the household-level, household covariates were collected in the following ways: reported variables (i.e. *presence* of a toilet at the household) were based on questions asked to the study participants during the baseline assessment visit, whilst observed variables (i.e. *type* of toilet), were answered by field teams observing the household infrastructure at multiple time points. These variables were screened for importance by a panel of environmental health specialists (Morse, Chidziwisano) with expertise on the risks and critical control points for faecal-oral transmission in Malawi [15,16,17].

Individual-level covariates such as age, gender, HIV status and antibiotic use were also selected. Antibiotic use was defined as the reported use of any antibiotics in the last six months (at baseline) and subsequently, for each follow-up visit, as the reported use of antibiotics between that visit and the previous one. We excluded long-term antibiotic therapy such as cotrimoxazole prophylactic therapy (CPT) in order to capture only the immediate change expected to occur following short courses of therapy. Household-level covariates such as household income and size were also included.

Stool samples from participants were cultured for growth of ESBL-producing bacteria on ESBL CHROMagar™ chromogenic agar (CHROMagar™, France). Bacterial colonies were classified by colour into categories, and speciation of blue colonies took place to identify ESBL-producing *K. pneumoniae* using polymerase chain reaction (PCR) as previously described in [14]. Throughout this paper, colonisation refers to asymptomatic carriage of either of these pathogens.

Household, individual and WASH datasets were merged together to form a unique dataset and all variables went through cleaning, removing incomplete and duplicate records assuming missingness at random.

Univariable logistic models were then used to investigate the effect of WASH infrastructure and associated behaviours on colonisation with either ESBL-producing *E. coli* or ESBL-producing *K. pneumoniae*. We started by looking at the effect of the study area on colonisation in order to separate its potential effect from the effects of other risk factors and then ran the other univariable models accordingly. This allowed for a refinement of the variables to be included in the full modelling framework, by retaining only the covariates which returned a p-value <0.2. For both the ESBL-producing *E. coli* and the ESBL-producing *K. pneumoniae* models, we pragmatically set this threshold to avoid missing the identification of important covariates.

### Multilevel harmonic hierarchical regression

The modelling framework is detailed in S1 Appendix. Briefly, we used a hierarchical multivariable harmonic logistic regression, incorporating covariates selected by the univariable analysis. In addition to the covariates, we included study region, annual and bi-annual harmonic terms to capture seasonality, and a serially-correlated household-level random effect to reflect temporal correlation in household-level prevalence between timepoints as well as heterogeneity in prevalence between households over and above that explained by the covariates. The analysis was carried out twice, for each of our *E. coli* and *K. pneumoniae* binary colonisation response variables.

A Bayesian approach to inference was used to allow for more flexibility in designing our model, using prior information from recent work on the dynamics of gut mucosal information with ESBL-producing Enterobacterales in Malawi [18] and to explore different forms of the temporal correlation structure at the household level. The model was fitted using the standard implementation of the No-U-Turn Sampler (NUTS) in the Stan modelling language [19] in R v4.1.1 through the package RStan [20]. The model was run with 20000 iterations for each of three independent Markov chains. Convergence was evaluated by inspection of trace plots and the Gelman-Rubin statistic being close to 1. Posterior estimates of parameters were expressed as medians with 95% credible intervals.

Initial model exploration considered adding the serially-correlated random effect at the individual-level, but found no evidence of individual-level temporal correlation. We therefore applied the serial correlation to a household-level random effect, enabling us to quantify both a household-effect (over and above that due to observed covariate information) and to account for possible “household contamination” with an ESBL as a result of a longer-term transmission process operating within the home. To ensure maximum identifiability of the random effects, we performed our analysis only on households with available sample results for all four time points. Throughout this analysis, the harmonic terms were always run as a single term. The variables were standardised, therefore the odds ratio should be interpreted as a change for each increase in standard deviation.

Ethical approval was obtained from Liverpool School of Tropical Medicine (LSTM) Research and Ethics Committee, UK (REC, #18-090) and College of Medicine Research and Ethics Committee, Malawi (#P.11/18/2541).

## Results

### Exploratory analysis

Twenty-six WASH variables were selected by the expert panel for further analysis. Among these variables, three were removed due to a lack of variation. Twenty-seven households were excluded due to missing enrolment or WASH data. Four individuals were removed due to missing data on age and gender, eighteen duplicate records were removed. Out of 2845 human stool samples collected by the field teams over time, forty-four duplicate records were removed. After merging, 224 samples were removed due to missing covariate data, and 84 samples from households ≥200 meters outside of the polygon limits, were also removed. This threshold permitted retention of households subsequently chosen by the field teams after refusal from the original sampled household. The combined dataset, which was used for the analysis, contained 2493 samples from 894 individuals in 259 households. The complete list of covariates and outcome variables can be found in S1 Table.

The distribution of samples, individuals, and households per polygon are presented in Table 1. Although the number of households and individuals in Ndirande and Chileka was higher than in Chikwawa, the number of available samples was similar with 36% in Chikwawa and Chileka and 28% in Ndirande.

**Table 1.** Distribution of the number of households, individuals and samples per polygon.

|  | **Ndirande** | **Chikwawa** | **Chileka** | **Total** |
| --- | --- | --- | --- | --- |
| **Households** | 96 | 64 | 99 | 259 |
| **Individuals** | 285 | 259 | 350 | 894 |
| **Samples** | 709 | 891 | 893 | 2493 |

After data cleaning, 233 individuals had one sample, 96 had two samples, 192 had three samples and 373 individuals had all four samples. The 129 households in which individuals had four available samples were used for the hierarchical model.

### Baseline participant data

Our age distribution data reflect Malawi’s population structure (Fig 2). People were considered adults at ≥16 years (54%, 483/894) and school age was ≥5 to 15 years (27.1%, 242/894), whilst 57.1% (510/894) participants were female with slightly varying proportions in each age group (65.2% (315/483) of adults and 46.7% (113/242) of school age children).

**Fig 1.**
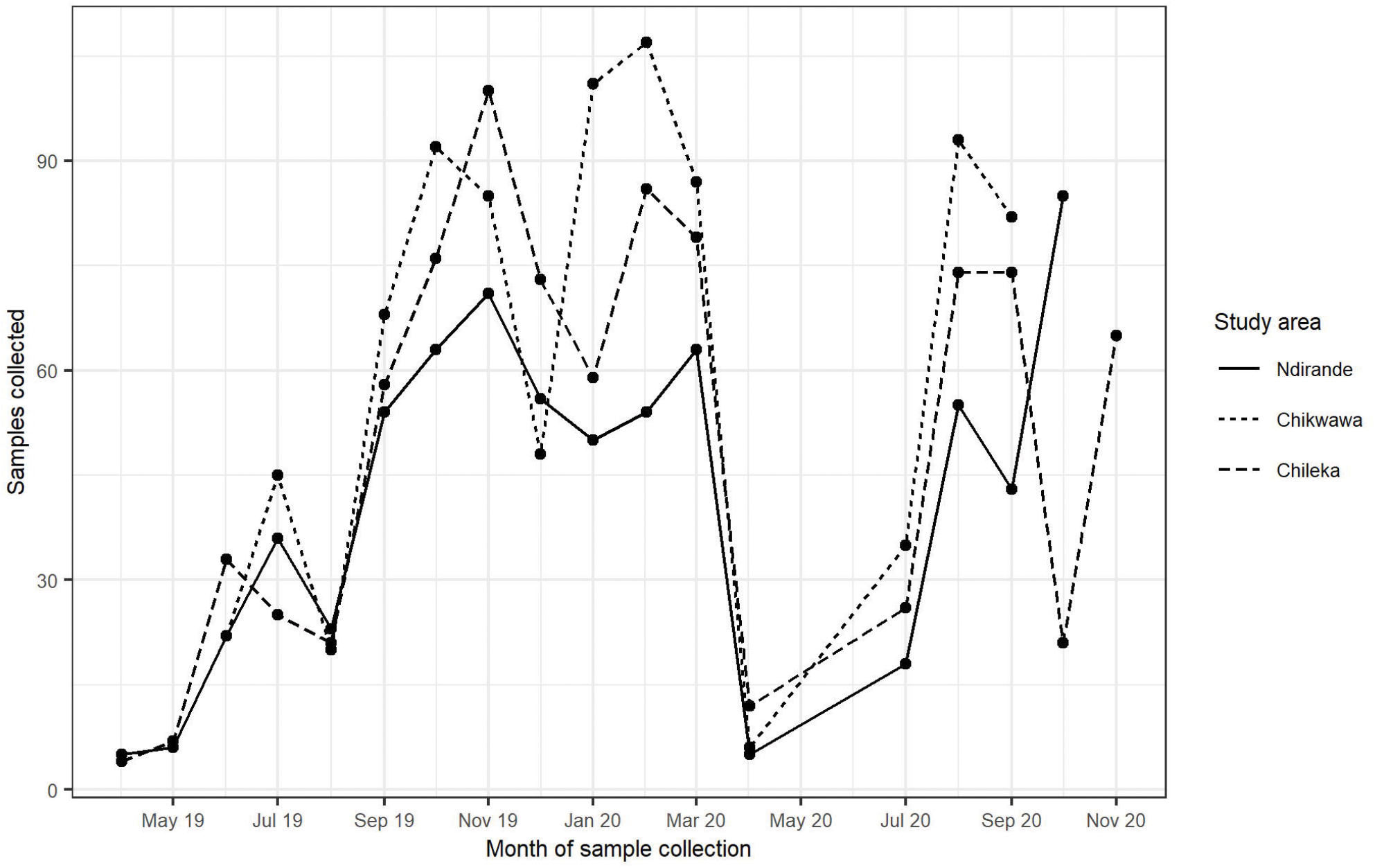
Distribution of samples collected, over time and by study area Baseline participant data.

**Fig 2.**
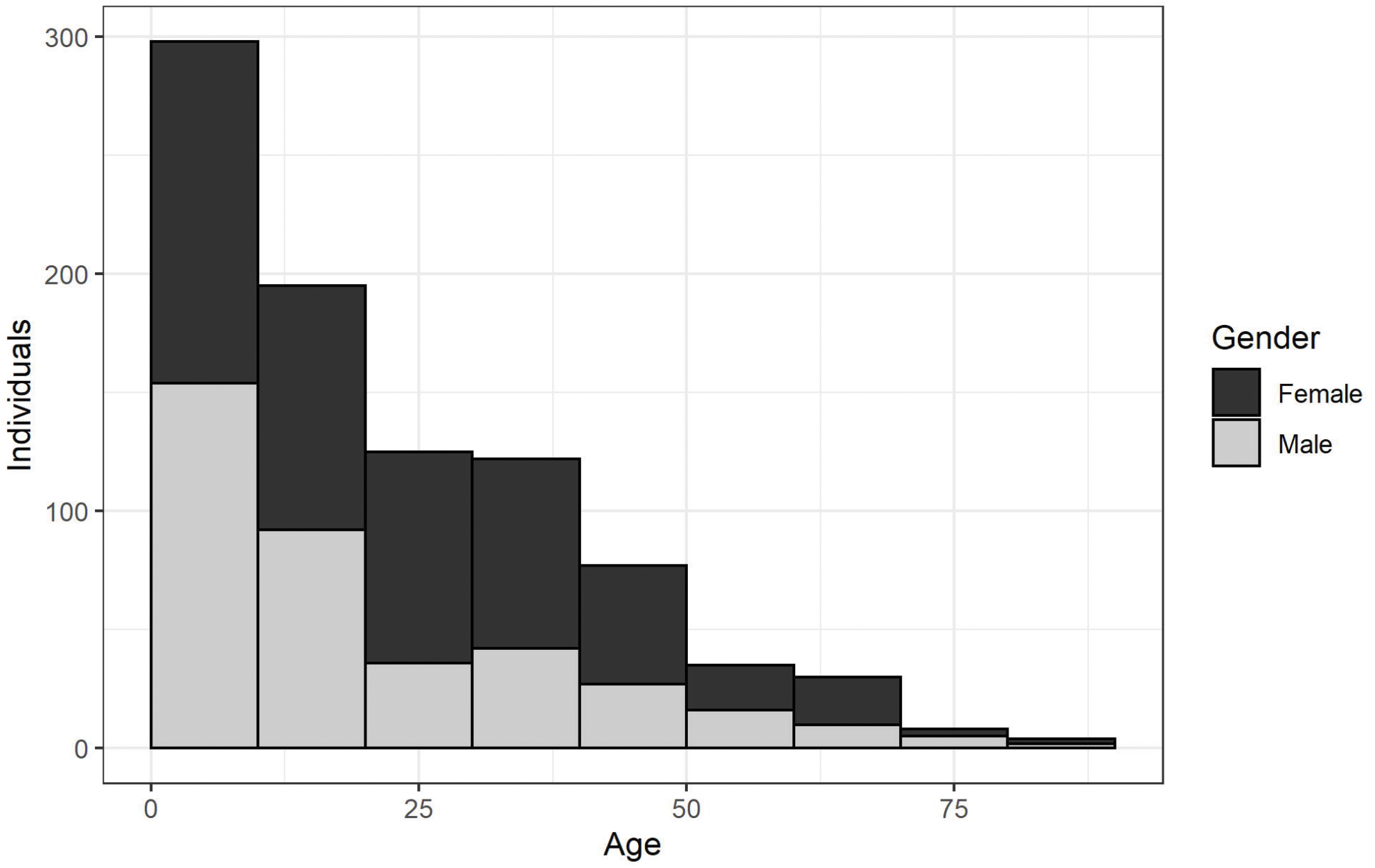
Distribution of age and gender for the 894 individuals.

At the first visit, 15.2% (129/851) of participants reported having taken at least one course of antibiotics in the preceding six months, while between subsequent visits, 6% (37/616), 9.4% (54/570) and 8.3% (38/456) reported at least one course of antibiotics. This varied by region, with 15.4% (137/891) in Chikwawa, 9.3% (66/709) in Ndirande and 6.2% (55/893) in Chileka.

Overall, the prevalence of ESBL-producing *E. coli* in our samples was 37% (922/2493) and the prevalence of ESBL-producing *K. pneumoniae* was 11.9% (296/2493). The prevalence of colonisation with ESBL-producing *E. coli* and ESBL-producing *K. pneumoniae* in our samples varied over time (Fig 3). The fluctuations in prevalence for both bacteria appear to follow a similar pattern in time. Some evidence of seasonality can be discerned for both, although more evidently for *E. coli*, with a decrease in prevalence during the dry season (May to October) followed by an increase during the wet season (November to April). This is followed by a sudden drop between April and July 2020, a period for which we have no available data due to Covid-19, and an increase in the last few months followed by a decrease in November 2020.

**Fig 3.**
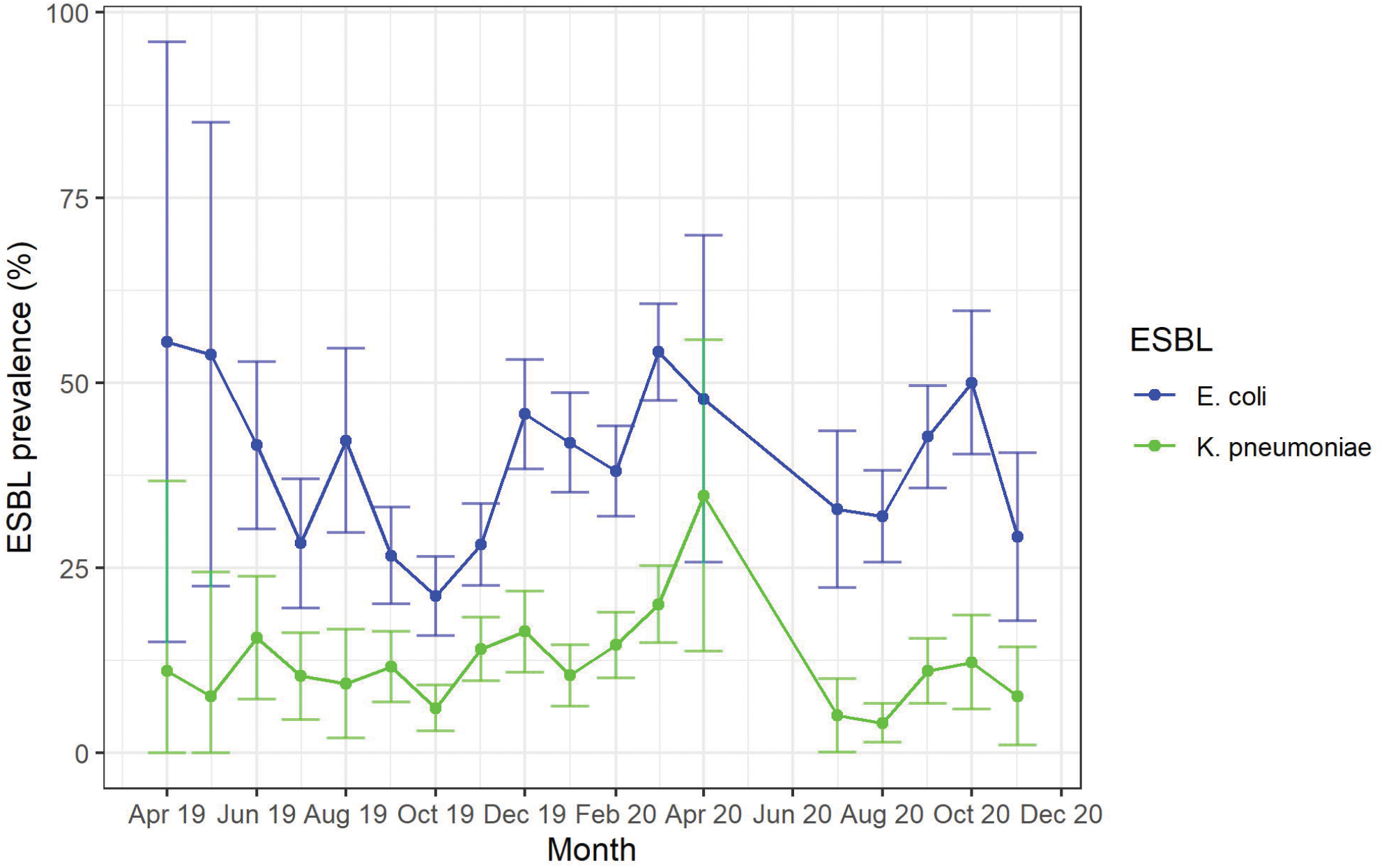
Prevalence of ESBL-producing *E. coli* and ESBL-producing *K. pneumoniae* per month.

### Baseline WASH data

Correlations between household-level WASH variables are depicted on the heatmap in Fig 4. There was a strong positive correlation between multiple animal-exposure related factors, for example bird owners appeared to be more likely to keep animals inside, therefore it was also more likely for these animals to come into contact with food preparation areas. Keeping animals inside also increased the likelihood of visible animal faeces around the household area. In terms of sanitation, the data suggested that with increasing income, there was increasing likelihood the household’s water drinking source came from a pipe, rather than a tube well or borehole. Moreover, increasing household income correlated with presence of hand washing facilities and soap presence in the household, and presence of cleaning materials such as toilet paper near the toilet. Some food consumption factors such as eating from shared plates appeared to be negatively correlated with the previously mentioned sanitation factors - the higher the income of the household is, the less chance individuals used shared plates when eating.

**Fig 4.**
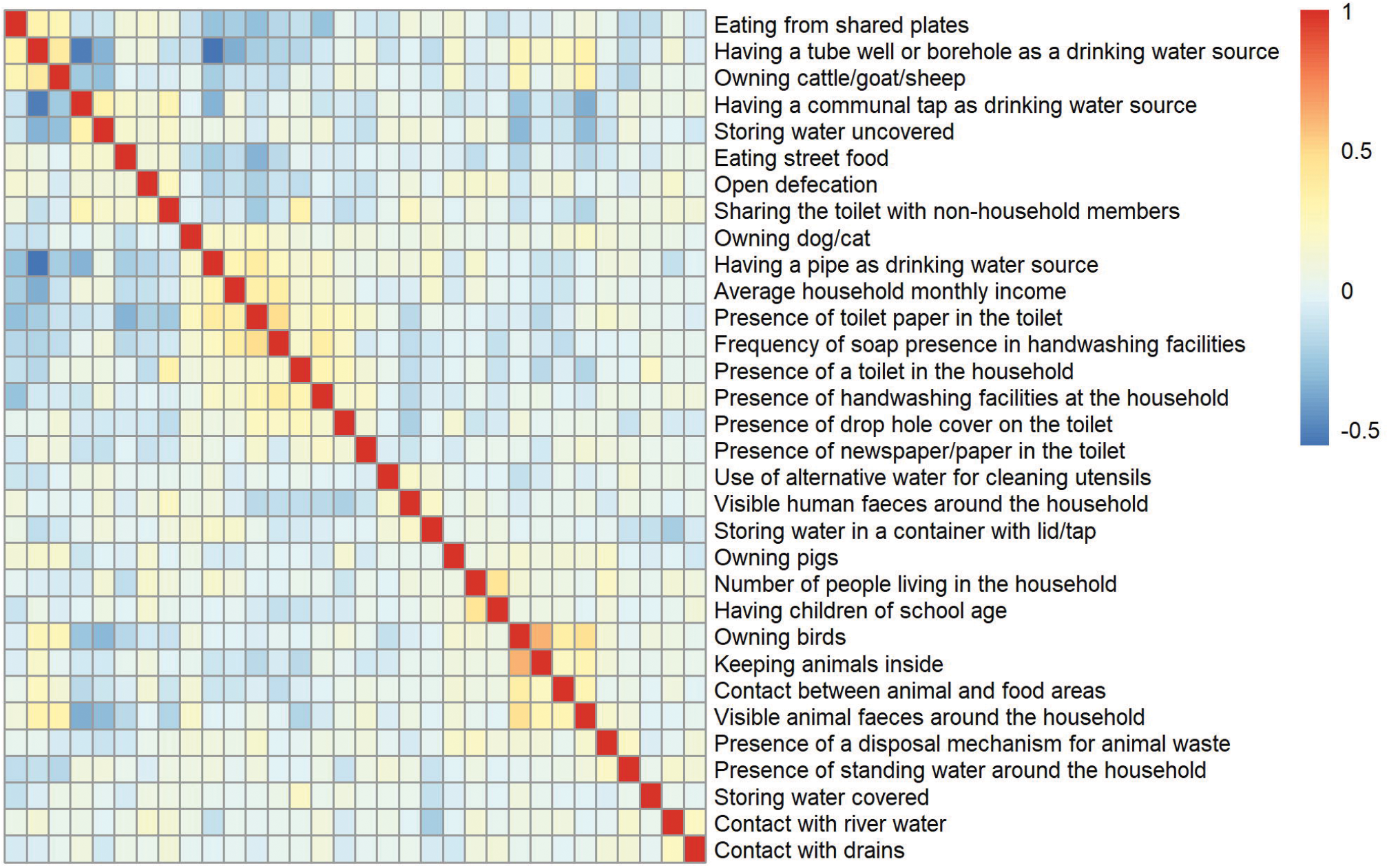
Correlation heatmap of household-level covariates.

### Impact of WASH infrastructure and associated behaviours on ESBL colonisation

In order to start exploring the effect of WASH variables on colonisation with either ESBL-producing *E. coli* or ESBL-producing *K. pneumoniae*, and to look at variability between regions, we first ran a generalised linear model including only the study area for both ESBL-producing *E. coli* and ESBL-producing *K. pneumoniae* (Table 2 and 3). We found a significant effect of the study area on ESBL-producing *E. coli*, with a higher risk of being colonised in Ndirande (OR 1.39 CI[1.13-1.70]) compared to Chileka. This was not the case for ESBL-producing *K. pneumoniae*, which showed no significant effect of the study area on the colonisation status. We observed that in the case of *E. coli*, WASH variables that vary depending on the study area the participant was in had a different signal if the study area was included as a variable in the analysis. Consequently, univariable analysis was run differently depending on the bacterial species with the study area included as a covariate when running univariable models for ESBL-producing *E. coli*, but not for ESBL-producing *K. pneumoniae*.

**Table 2.**
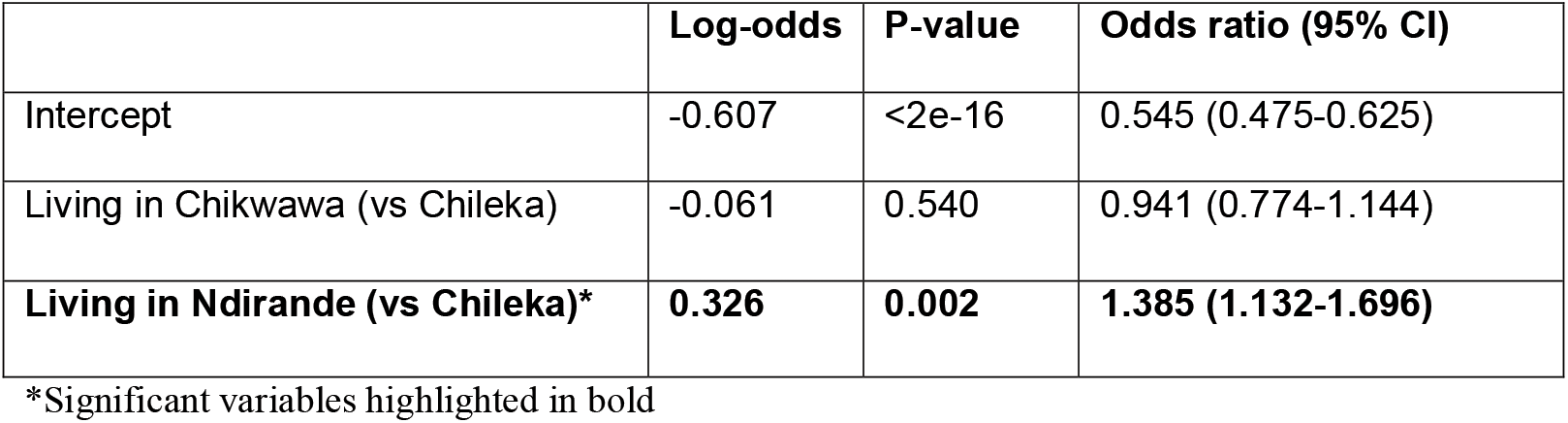
Relationship between the study area and the ESBL-producing *E. coli* colonisation status.

|  | Log-odds | P-value | Odds ratio (95% CI) |
| --- | --- | --- | --- |
| Intercept | -0.607 | <2e-16 | 0.545 (0.475-0.625) |
| Living in Chikwawa (vs Chileka) | -0.061 | 0.540 | 0.941 (0.774-1.144) |
| **Living in Ndirande (vs Chileka)\*** | **0.326** | **0.002** | **1.385 (1.132-1.696)** |

\*Significant variables highlighted in bold

**Table 3.** Relationship between the study area and the ESBL-producing *K. pneumoniae* colonisation status.

|  | Log-odds | P-value | Odds ratio (95% CI) |
| --- | --- | --- | --- |
| Intercept | -2.105 | <2e-16 | 0.122 (0.099-0.151) |
| Living in Chikwawa (vs Chileka) | 0.196 | 0.183 | 1.216 (0.912-1.622) |
| Living in Ndirande (vs Chileka) | 0.098 | 0.536 | 1.103 (0.809-1.504) |

### Human gut mucosal colonisation with ESBL-producing *E. coli*

Results from the univariable models showed that having a drinking water source coming from a tube well or a borehole, having a drop hole cover on the toilet and animals being able to enter into contact with the food areas all appeared to be highly significant (S2 Table). Whilst a positive association with ESBL-producing *E. coli* colonisation status was detected for the drinking water source coming from a tube well or a borehole, the opposite can be said for piped drinking water. This was consistent with the negative correlation between those two water variables noted previously in the correlation heatmap. Animal contact with food areas was positively associated with ESBL-*E. coli* colonisation, whilst having a drop hole cover on the toilet appeared to have a protective effect, being negatively associated with colonisation status. Additionally, variables such as keeping animals inside the house, having a toilet floor surfaced with soil and having clean paper in the toilet were also highly significant. Having clean paper in the toilet was negatively associated with colonisation status while the others showed a positive association. Other variables such as older age, the presence of open defecation in the area, owning cattle, sheep or goats, entering into contact with river water all were significant (<0.05) and were positively associated with the colonisation status. In contrast, male sex, higher income, having a disposal mechanism for animal waste, having a piped water drinking source, storing water in a container with lid and tap were all significantly negatively associated with ESBL-*E. coli* colonisation status (S2 Table).

We subsequently ran the hierarchical model. We found that men are less at risk of becoming colonised with ESBL-producing *E. coli* (OR 0.786 CrI[0.678-0.910]) and that having a tube well or a borehole as a water drinking source highly increases your risk of becoming colonised (OR 1.550 CrI[1.003-2.394]). Coming into contact with standing water also appeared to be negatively associated with colonisation status (OR 0.749 CrI[0.574-0.978]). Finally, there was an apparent signal of annual seasonality noticeable from the presence of part of the harmonic term (Table 4).

**Table 4.** Temporal model results for ESBL-producing *E. coli* colonisation status.

|  | Log-odds | Odds ratio (95% CrI) |
| --- | --- | --- |
| (Intercept) | -0.716 | 0.489 (0.360-0.663) |
| Reactive to HIV testing (vs non-reactive) | 0.027 | 1.027 (0.863-1.223) |
| Unknown HIV status (vs non-reactive) | -0.031 | 0.969 (0.808-1.163) |
| Recent use of antibiotics | 0.093 | 1.097 (0.946-1.274) |
| Age | 0.132 | 1.141 (0.970-1.343) |
| **Being male (vs female)\*** | **-0.241** | **0.786 (0.678-0.910)** |
| Average household monthly income | 0.226 | 1.254 (0.916-1.715) |
| Open defecation | 0.054 | 1.055 (0.826-1.349) |
| Presence of a disposal mechanism for animal waste | 0.104 | 1.110 (0.857-1.437) |
| Eating from shared plates | -0.245 | 0.783 (0.598-1.024) |
| Having a pipe as drinking water source | 0.132 | 1.141 (0.818-1.592) |
| **Having a tube well/ borehole as drinking water source** | **0.438** | **1.550 (1.003-2.394)** |
| Use of alternative water for cleaning utensils | 0.014 | 1.014 (0.802-1.283) |
| Owning cattle, goats or sheep | 0.139 | 1.149 (0.892-1.480) |
| Keeping animals inside | 0.075 | 1.078 (0.852-1.364) |
| Contact with river water | 0.048 | 1.049 (0.791-1.391) |
| Toilet floor material: none (vs concrete/wood) | 0.123 | 1.131 (0.799-1.600) |
| Toilet floor material: soil (vs concrete/wood) | 0.131 | 1.140 (0.820-1.585) |
| Presence of drop hole cover on the toilet | -0.202 | 0.817 (0.626-1.067) |
| Presence of newspaper/paper in the toilet | -0.155 | 0.856 (0.670-1.094) |
| Frequency of soap presence in handwashing facilities | -0.000 | 1.000 (0.640-1.563) |
| Storing water covered | -0.179 | 0.836 (0.537-1.302) |
| Storing water in a container with lid/tap | -0.034 | 0.967 (0.754-1.240) |
| Contact between animal and food areas | 0.218 | 1.244 (0.983-1.573) |
| **Presence of standing water around the household** | **-0.289** | **0.749 (0.574-0.978)** |
| Number of days since the first sample | 0.167 | 1.182 (0.869-1.608) |
| **Harmonic term (sinday)** | **-0.466** | **0.628 (0.453-0.869)** |
| Harmonic term (cosday) | 0.371 | 1.449 (0.958-2.191) |
| Harmonic term (sinday2) | -0.084 | 0.919 (0.644-1.314) |
| Harmonic term (cosday2) | 0.018 | 1.018 (0.711-1.457) |
| Living in Chikwawa (vs Chileka) | -0.276 | 0.759 (0.527-1.093) |
| Living in Ndirande (vs Chileka) | 0.386 | 1.471 (0.980-2.207) |

\*Significant variables highlighted in bold

Using the covariance structure (S1 Appendix), we found a range of temporal correlation estimated at 77.85 days (CrI [30.85-140.60]), thus samples that have been sampled in the same household more than 77 days apart are effectively uncorrelated.

Parameter estimates are shown in S3 Table. The densities of the priors and posteriors of all three parameters can be found in S1 Fig. Visual inspection of the trace plots in S2 Fig and calculations of the Gelman-Rubin statistic resulting close to 1 for all parameter estimates indicates that the model has fitted properly.

### Human gut mucosal colonisation with ESBL-producing *K. pneumoniae*

In the case of ESBL-producing *K. pneumoniae*, univariable results showed that the household size was the only highly significant variable (p<0.01) except for the harmonic terms, revealing that with increasing household size, there was greater the risk of being colonised with ESBL-*K. pneumoniae*. Variables such as eating street food, eating from shared plates, owning birds and coming into contact with drains were also significant. Eating street food and eating from shared plates surprisingly appeared to have a protective effect on the ESBL-producing *K. pneumoniae* colonisation status. Owning birds and coming into contact with drains were both positively associated with colonisation status (S4 Table).

The hierarchical model for ESBL *K. pneumoniae* found that having previously used antibiotics (in the last six months or in-between visits) increased the risk of being colonised with ESBL-producing *K. pneumoniae* (OR 1.281 CrI[1.049-1.565]). We also saw a negative association between eating from shared plates and colonisation (OR 0.672 CrI[0.460-0.980]). Finally, there was a signal of annual seasonality noticeable from the presence of part of the harmonic term, similar to the one we found for ESBL *E. coli*. These results are presented in Table 5.

**Table 5.**
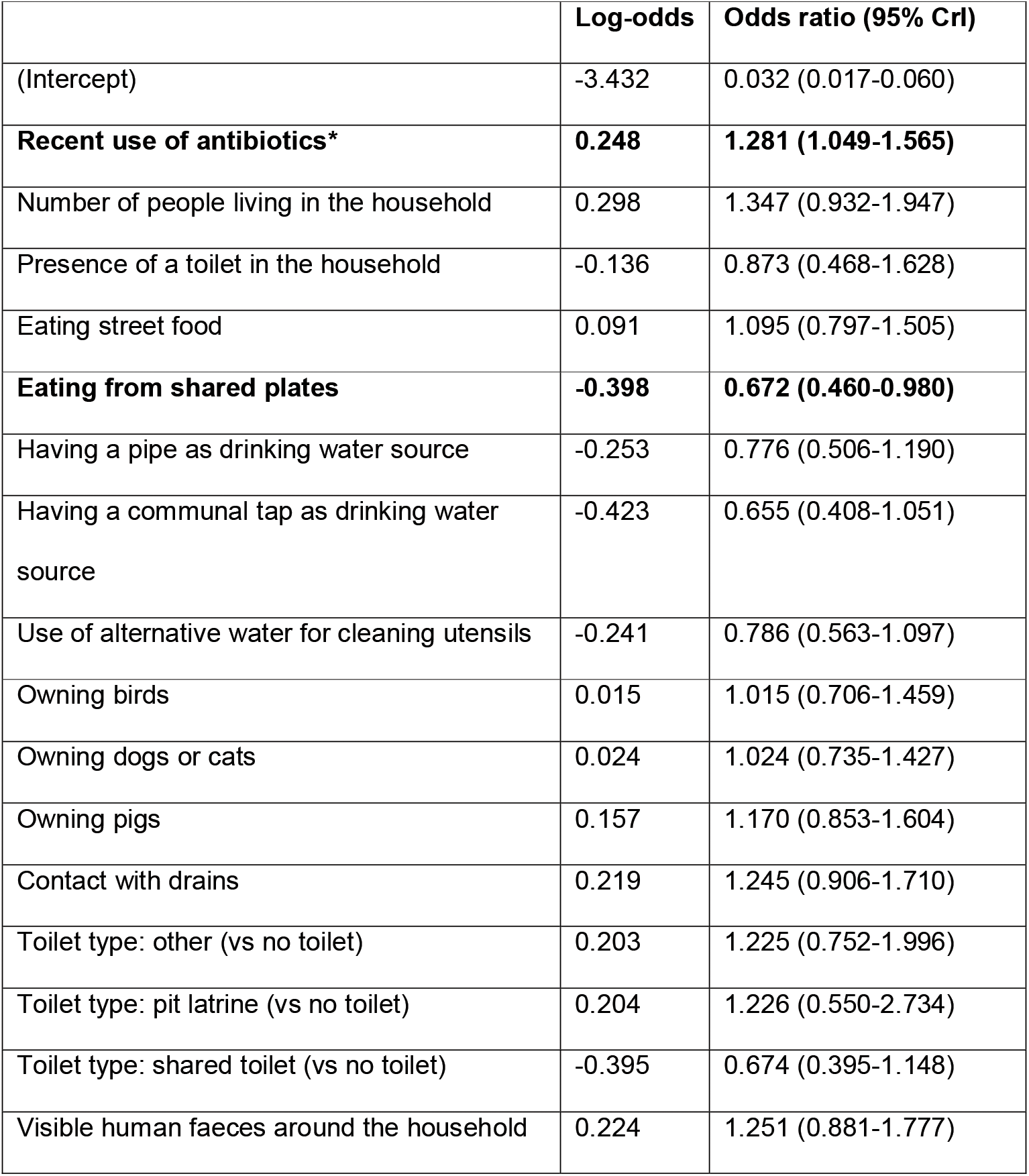
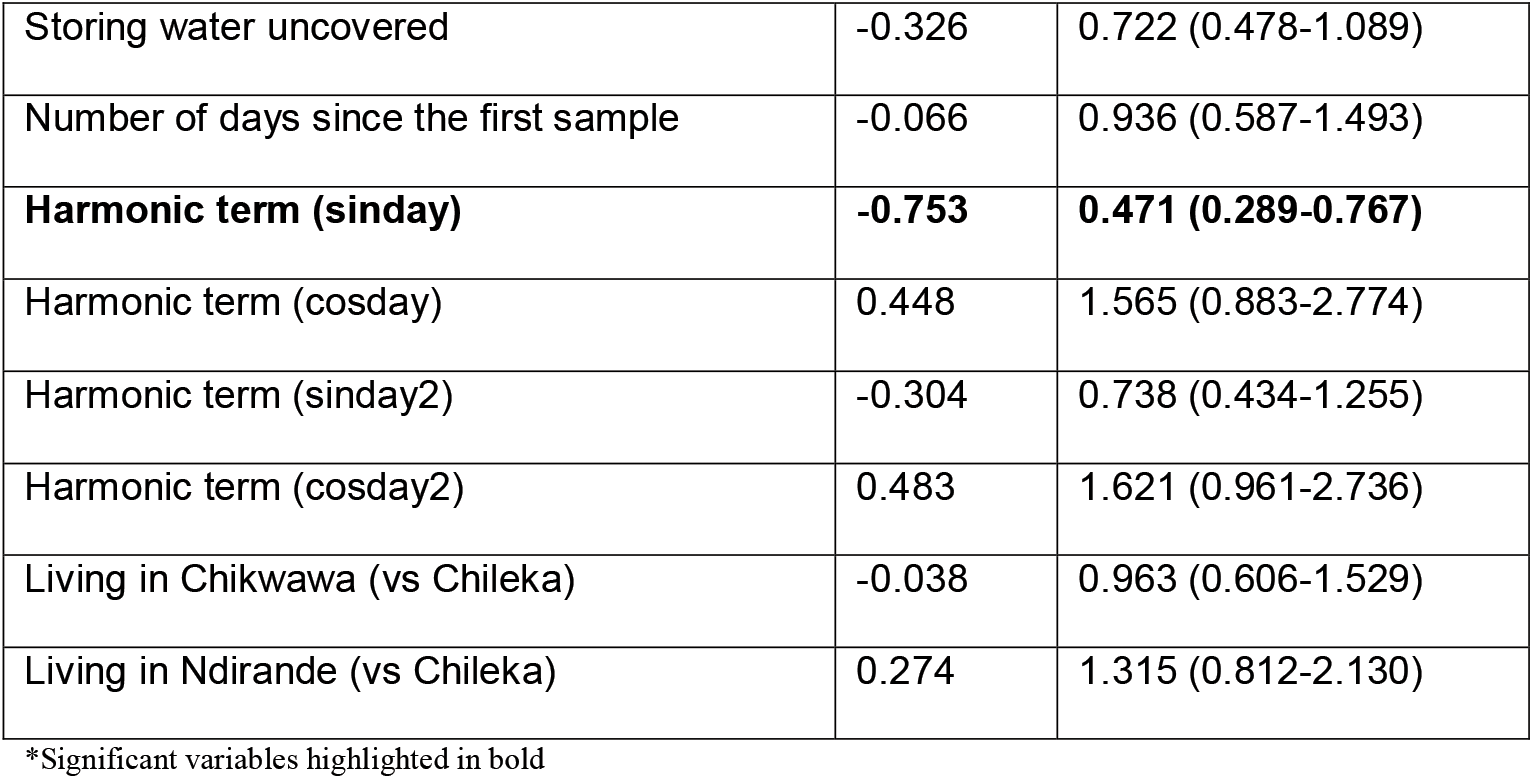
Temporal model results for ESBL-producing *K. pneumoniae* colonisation status.

|  | Log-odds | Odds ratio (95% CrI) |
| --- | --- | --- |
| (Intercept) | -3.432 | 0.032 (0.017-0.060) |
| **Recent use of antibiotics\*** | **0.248** | **1.281 (1.049-1.565)** |
| Number of people living in the household | 0.298 | 1.347 (0.932-1.947) |
| Presence of a toilet in the household | -0.136 | 0.873 (0.468-1.628) |
| Eating street food | 0.091 | 1.095 (0.797-1.505) |
| **Eating from shared plates** | **-0.398** | **0.672 (0.460-0.980)** |
| Having a pipe as drinking water source | -0.253 | 0.776 (0.506-1.190) |
| Having a communal tap as drinking water source | -0.423 | 0.655 (0.408-1.051) |
| Use of alternative water for cleaning utensils | -0.241 | 0.786 (0.563-1.097) |
| Owning birds | 0.015 | 1.015 (0.706-1.459) |
| Owning dogs or cats | 0.024 | 1.024 (0.735-1.427) |
| Owning pigs | 0.157 | 1.170 (0.853-1.604) |
| Contact with drains | 0.219 | 1.245 (0.906-1.710) |
| Toilet type: other (vs no toilet) | 0.203 | 1.225 (0.752-1.996) |
| Toilet type: pit latrine (vs no toilet) | 0.204 | 1.226 (0.550-2.734) |
| Toilet type: shared toilet (vs no toilet) | -0.395 | 0.674 (0.395-1.148) |
| Visible human faeces around the household | 0.224 | 1.251 (0.881-1.777) |
| Storing water uncovered | -0.326 | 0.722 (0.478-1.089) |
| Number of days since the first sample | -0.066 | 0.936 (0.587-1.493) |
| **Harmonic term (sinday)** | **-0.753** | **0.471 (0.289-0.767)** |
| Harmonic term (cosday) | 0.448 | 1.565 (0.883-2.774) |
| Harmonic term (sinday2) | -0.304 | 0.738 (0.434-1.255) |
| Harmonic term (cosday2) | 0.483 | 1.621 (0.961-2.736) |
| Living in Chikwawa (vs Chileka) | -0.038 | 0.963 (0.606-1.529) |
| Living in Ndirande (vs Chileka) | 0.274 | 1.315 (0.812-2.130) |

\*Significant variables highlighted in bold

We found a range of temporal correlation for ESBL-*K. pneumoniae* colonisation estimated at 54.29 days (CrI [12.91-130.43]), thus samples that have been sampled in the same household more than 54 days apart are effectively uncorrelated. Parameter estimates are shown in S5 Table. The densities of the priors and posteriors of all three parameters can be found in S3 Fig. Convergence was verified by looking at the trace plots in S4 Fig and we confirmed that the Gelman-Rubin statistic was close to 1 for all parameter estimates.

## Discussion

This study identified varying prevalence of ESBL colonisation over time for both ESBL-producing *E. coli* and ESBL-producing *K. pneumoniae*. A decrease in prevalence was observed during the dry season, followed by an increase during the wet season, and this apparent seasonality was confirmed by the model results. Potential explanations for this variation include the accumulation of mud and floodwater due to the heavy rain in the wet season, which might lead to more contact between individuals and contaminated soil or water.

Additionally, the increase in time spent indoors when heavy rain occurs might lead to higher within-household transmission.

The correlation heatmap (Fig 4) suggested that the socioeconomic status of the household greatly influences the WASH situation of the household, and that higher income allows for a better access to cleaner water and easier availability of sanitation and hygiene products. In particular, there was a positive association between using a tube well or a borehole as a drinking water source and being ESBL-colonised, which is confirmed in the temporal model for ESBL-producing *E. coli*. Being female was also identified as a risk factor for ESBL-producing *E. coli*, which could be explained by the fact that traditionally women are more likely to perform domestic duties -- such as laundry, housework, and childcare -- which would place them at higher risk of being in contact with the faecally-contaminated environment. However, no direct association was found between income and gut mucosal colonisation in the model. Further work is needed to understand the association between income and gut mucosal colonisation, but it is noteworthy that the vast majority of households in the study were below the World Bank defined threshold of absolute poverty (<$1.90/day per individual) and that income alone is a poor indicator of wealth.

Other variables identified by the univariable models as conferring a highly significant increased risk of being colonised with ESBL-producing *E. coli* included the study area and permitting animals inside the home, and allowing them to contact food preparation areas. This is common practice in low- and middle-income countries (LMICs), where animal husbandry is frequently a primary source of income [21]. However, this practice increases the risk of faecal contamination of the soil by enteric pathogens like *E. coli* [22] and therefore puts household members, especially young children, at higher risk for exposure to faecal pathogens and enteric infections [23]. We also found that having access to cleaning materials such as paper in the toilet and a drop hole cover on the toilet were both negatively associated with ESBL-*E. coli* colonisation. Such infrastructure is used to prevent flies from accessing faecal matter, thus this association is consistent with studies which have shown the role of flies in transporting and transmitting *E. coli* [24,25].

Antibiotic use was identified as a risk factor for carriage of ESBL-producing *K. pneumoniae*, which is consistent with previous studies in sub-Saharan Africa [5,8,9,10]. The highest level of antibiotic use reported in time was at the baseline visit, with lower rates reported at subsequent visits, likely due to shorter intervals between subsequent visits compared to the initial six month question. The reason for the regional variation is uncertain. Chikwawa being the rural area of the study [14], participants may have encountered more often organisations such as non-governmental organisations that might have been able to offer them treatment or antibiotics, or participants might have had increased access to antibiotics due to the greater presence of animal farming [26]. This emphasizes the importance of antimicrobial exposure in driving ESBL colonisation, thus highlighting the need for a more responsible antibiotic consumption.

Given our model and dataset, eating from shared plates rather than from separate plates as well as the presence of standing water around the household appeared to have a protective effect. This is a surprising result, since plate-sharing, which is common in LMICs [27] and has been associated with other enteric pathogens in other settings [28], would seem to promote transmission between individuals. Similarly, since wastewater is known to play a role in the transmission of AMR [12], the presence of standing water around the household would also seem to promote transmission. We caution that these results might represent a Type I error (in a Bayesian context), and that further research into WASH behavioural patterns and interactions with other explanatory variables would be necessary to confirm or refute our findings.

For ESBL-producing *K. pneumoniae*, at the univariable level, household size was the only highly significant risk factor, highlighting the importance of the household in driving ESBL transmission. Other variables identified as conferring a significant increased risk of being colonised with ESBL-producing *K. pneumoniae* included owning birds, which are known to be responsible for faecal contamination of the household environment in LMICs [29], and entering into contact with drains, highlighting again the importance of interactions between animals, humans and the environment.

The temporal models for both bacterial species detected a temporal correlation range of seven to ten weeks. In other words, two samples taken within that time frame are more likely to both be colonised than if spread apart in time any further. Though our method is designed to only detect association between ESBL prevalence in subsequent follow-ups, this does suggest that within-household transmission occurs within this time frame. Subsequent causal inference studies would, however, be required to confirm this.

This study had some limitations. The volume of information available from various questionnaires was considerable and for that reason, we had to pre-select variables based on their perceived importance by environmental health experts. Although we found temporal correlation at household-level, we could not find any at individual-level, which suggested that an individual’s samples could be seen as independent from each other. Potential explanations for this lack of temporal correlation at the individual level include the use of stool samples over rectal swabs, which may have been better for screening. Additionally, the laboratory protocol for testing was qualitative, discriminating only between presence or absence of ESBLs without quantification. Further work is needed to consider the impact of microbiological methods on informing these models (i.e. time in enrichment broth and/or quantification by minimal probable number estimates). Whole genome sequencing will allow for a more precise investigation of the samples to get a better understanding of the linkage between sequence types. The COVID-19 pandemic also played a role in derailing the microbiological sampling for our study and potentially impacting our results. The pandemic caused the sampling and microbiological testing to be suspended between April and July 2020, which caused some delay in our data collection.

Our study suggests that WASH factors and environmental hygiene are key drivers of AMR-transmission in Malawi, consistent with findings in other African settings [30]. Our results also point towards acquisition of ESBL-producing *E. coli* through contaminated water and/or inappropriate WASH infrastructure. Additionally, seasonality and gender also suggest the importance of environmental hygiene and practices in driving ESBL-producing *E. coli* transmission. This underlines the need for improved access to clean water and suggests that associating WASH behavioural practice with better WASH conditions would be instrumental in decreasing transmission. However, for ESBL-producing *K. pneumoniae*, previous antibiotic use was identified as a risk factor, therefore emphasizing the importance of antimicrobial exposure in driving ESBL-producing *K. pneumoniae* transmission and the need for improved infection prevention and control (IPC) measures and antibiotic usage and stewardship training. A better understanding of how the WASH conditions of the different communities impacts ESBL colonisation and transmission will inform public health responses to the challenge presented by AMR and enable design of effective intervention strategies in Southern and Eastern Africa.

## Data Availability

All data produced in the present work are contained in the supplementary material.

## Acknowledgments

We would like to thank the participants for taking part in the study, as well as the wider DRUM consortium (https://www.drumconsortium.org/about/the-drum-team) for their advice and support. We thank Lancaster University for the use of their High End Computer, and in particular Dr Mike Pacey for his help in setting up our software stack.

For the purpose of open access, the author has applied a CC BY public copyright licence (where permitted by UKRI, ‘Open Government Licence’ or ‘CC BY-ND public copyright licence may be stated instead) to any Author Accepted Manuscript version arising.

## Data availability statement

The authors confirm that the data supporting the findings of this study are available within its supplementary materials.

## Supplementary information

**S1 Appendix. Modelling framework**.

**S1 Dataset. Dataset used for the modelling.**

**S1 Table. Covariates and outcome variables**.

**S2 Table. Univariable analysis results between ESBL-producing *E. coli* colonisation status and each variable accounting for the study area**.

**S3 Table. Estimates for φ, σ and τ in the ESBL-*E. coli* temporal model**.

**S4 Table. Univariable analysis results between ESBL-producing *K. pneumoniae***

**colonisation status and each variable**.

**S5 Table. Estimates for φ, σ and τ in the ESBL-*K. pneumoniae* temporal model**.

**S1 Fig. Prior and posterior density of φ, σ and τ (left to right, without warm-up) for the ESBL *E. coli* temporal model**.

**S2 Fig. Trace plots of φ, σ and τ (left to right, without warm-up) for the temporal model for ESBL *E. coli***.

**S3 Fig. Prior and posterior density of φ, σ and τ (left to right, without warm-up) for the ESBL *K. pneumoniae* temporal model**.

**S4 Fig. Trace plots of φ, σ and τ (left to right, without warm-up) for the temporal model for ESBL *K. pneumoniae***.

