## Supplementary Appendix for "Risk factors, temporal dependence, and seasonality of human ESBL-producing *E. coli* and *K. pneumoniae* colonisation in Malawi: a longitudinal model-based approach"

**S1 Appendix: Modelling framework**

Let $y_{ijt}$ be 1 if the individual $i$ tested at household $j$ at time $t$ is colonised, our model can be expressed as:

$$logit\left( p_{ijt} \right)=\alpha+\left( x_{ij} \right)^{T}\beta+\theta_{1}cos\left( \frac{2\pi t}{365T} \right)+\theta_{2}sin\left( \frac{2\pi t}{365T} \right)+\theta_{3}cos\left( \frac{2\pi t}{365T} \right)+\theta_{4}sin\left( \frac{2\pi t}{365T} \right)+u_{jt}$$

where $Y_{ijt}$follows a Bernoulli distribution with probability $p_{ijt}$ i.e. $Y_{ijt}\sim\text{Bernoulli}\left( p_{ijt} \right)$, $\alpha$ is the intercept, $x_{ij}$ are the household-level and individual-level explanatory variables, $\beta$ are the regression coefficients for the fixed effects, $\theta_{1}\text{cos}\left( \frac{2\pi t}{365T} \right)+\theta_{2}\text{sin}\left( \frac{2\pi t}{365T} \right)+\theta_{3}\text{cos}\left( \frac{2\pi t}{365T} \right)+\theta_{4}\text{sin}\left( \frac{2\pi t}{365T} \right)$ the annual and bi-annual harmonic terms, with $T=\left( 1,\frac{1}{2} \right)$ the period in years. In order to look at the temporal correlation between different time points, we included a temporally correlated random effect $u_{jt}$ at the household-level with covariance structure:

$$\text{cov}\left( u_{jt},u_{jt+s} \right)=\sigma^{2}e^{-\frac{s^{2}}{\phi^{2}}}+\tau^{2}$$

where $s$ is the distance (in time) between two time points, $\phi$ is the scale of temporal correlation, $\sigma^{2}$ is the variance of the temporal process, $\tau^{2}$ is the nugget effect. Prior distributions were chosen as follows:

$$\alpha\sim\text{Normal}\left( 0,100 \right) \beta\sim\text{Normal}\left( 0,10 \right) \sigma^{2}\sim\text{Gamma}\left( 2,1 \right)$$

$$\tau\sim\text{Gamma}\left( 2,1 \right) \phi\sim\text{Gamma}\left( 4,0.125 \right)$$

The prior distribution for $\phi$ was based on recent work on the dynamics of gut mucosal colonisation with ESBL-producing Enterobacteriaceae in Malawi where they found an estimated mean time of 43 days between the ESBL-E colonised and uncolonised states [18].
