## Supplementary Table 1 for "Risk factors, temporal dependence, and seasonality of human ESBL-producing *E. coli* and *K. pneumoniae* colonisation in Malawi: a longitudinal model-based approach"

|  | | | **Variable** | **Description** | **S1 Dataset name** |
| --- | --- | --- | --- | --- | --- |
| **Individual factors** | | | HIV status | Categorical, {Reactive/Unknown/Non-reactive} (Reference: Non-reactive) | ip_hivstatus |
|  |  |  | Recent use of antibiotics | Binary, {1 = at least a course of antibiotic taken in the last 6 months or in-between visits, 0 = none} | antibio |
|  |  |  | Age | Continuous, age at enrolment | age |
|  |  |  | Being male (vs female) | Categorical, {Male/Female} (Reference: Female) | ip_sex |
| **Household factors** | | | Number of people living in the household | Continuous, number of people living in the household at baseline | hhcount |
|  |  |  | Average household monthly income | Continuous, average monthly household income (MWK) at baseline | hh_income |
|  |  |  | Having children of school age | Binary, {1 = children of school age living in the household, 0 = no children of school age living in the household} | schoolch |
| **WASH factors** | **Reported** | **Sanitation factors** | Presence of a toilet in the household | Binary, {1 = toilet present, 0 = toilet absent} | hh_toilet |
|  |  |  | Open defecation | Binary, {1 = open defecation reported, 0 = no open defecation reported} | hh_opendefecate |
|  |  |  | Sharing the toilet with non-household members | Binary, {1 = share toilet, 0 = do not share toilet} | hh_toiletshare |
|  |  |  | Presence of a disposal mechanism for animal waste | Binary, {1 = disposal mechanism available, 0 = no disposal mechanism available} | hh_disposal |
|  |  | **Food factors** | Eating street food | Binary, {1 = eat street food at times, 0 = never eat street food} | hh_food10 |
|  |  |  | Eating from shared plates | Binary, {1 = shared plates used, 0 = separate plates used} | hh_eating |
|  |  | **Water factors** | Having a pipe as drinking water source | Binary, {1 = yes, 0 = no} | hh_pipewater |
|  |  |  | Having a communal tap as drinking water source | Binary, {1 = yes, 0 = no} | hh_tapwater |
|  |  |  | Having a tube well/borehole as drinking water source | Binary, {1 = yes, 0 = no} | hh_tubewellwater |
|  |  |  | Use of alternative water for cleaning utensils | Binary, {1 = use of different water than the one used for drinking, 0 = use of same water than the one used for drinking} | hh_utensilwater |
|  |  | **Animal factors** | Owning birds | Binary, {1 = yes (owns one or more), 0 = no} | hh_birds |
|  |  |  | Owning cattle, goats or sheep | Binary, {1 = yes (owns one or more), 0 = no} | hh_cgs |
|  |  |  | Owning dogs or cats | Binary, {1 = yes (owns one or more), 0 = no} | hh_dogcat |
|  |  |  | Owning pigs | Binary, {1 = yes (owns one or more), 0 = no} | hh_pigs |
|  |  |  | Keeping animals inside | Binary, {1 = yes, 0 = no} | hh_aninside |
|  |  | **Broader environment factors** | Contact with river water | Binary, {1 = any adult or child at the household interact with river water, 0 = no adult or child at the household interact with river water} | hh_riverwater |
|  |  |  | Contact with drains | Binary, {1 = any adult or child at the household interact with drains, 0 = no adult or child at the household interact with drains} | hh_drains |
|  | **Observed** | **Sanitation factors** | Toilet type | Categorical, {Pit latrine/Shared toilet/No toilet/Other} (Reference: No toilet) | hh_toilettype |
|  |  |  | Toilet floor material | Categorical, {Concrete or wood/Soil/No toilet} (Reference: Concrete or wood) | hh_toiletfloor |
|  |  |  | Having a drop hole cover on the toilet | Binary, {1 = drop hole cover present, 0 = drop hole cover absent} | hh_drophole |
|  |  |  | Presence of toilet paper in the toilet | Binary, {1 = toilet paper present, 0 = toilet paper absent} | cleantp |
|  |  |  | Presence of newspaper/paper in the toilet | Binary, {1 = newspaper or paper present, 0 = newspaper or paper absent} | cleanp |
|  |  |  | Visible human faeces around the household | Binary, {1 = visible human stool, 0 = no visible human stool} | humanf |
|  |  | **Hygiene factors** | Presence of handwashing facilities (hwf) in the household | Binary, {1 = present anywhere within the household, 0 = absence within the household} | toif |
|  |  |  | Frequency of soap presence in handwashing facilities | Continuous, number of hwf with soap in the house over total number of hwf present in the house | soapiness |
|  |  | **Water factors** | Storing water covered | Binary, {1 = water stored at the house covered, 0 = no water stored at the house covered} | hh_covstwater |
|  |  |  | Storing water uncovered | Binary, {1 = water stored at the house uncovered, 0 = no water stored at the house uncovered} | hh_uncovstwater |
|  |  |  | Storing water in a container with lid/tap | Binary, {1 = water stored at the house in a container with lid and/or tap, 0 = no water stored at the house in a container with lid and/or tap} | hh_latstwater |
|  |  | **Animal factors** | Contact between animals and food areas | Binary, {1 = animal seen in contact with food areas, 0 = no animal seen in contact with food areas} | contactfood |
|  |  |  | Visible animal faeces around the household | Binary, {1 = animal faeces seen around the household, 0 = no animal faeces seen around the household} | hh_anfab |
|  |  | **Broader environmental factors** | Presence of standing water around the household | Binary, {1 = standing water seen, 0 = standing water not seen} | hh_stdgwater |
| **Temporal factors** | | | Number of days since the first sample | Continuous, number of days since the first sample was taken | diffdate |
|  |  |  | Harmonic terms (sinday, cosday, sinday2, cosday2) | Continuous, described in the modelling framework (S1 Appendix) | sinday  cosday  sinday2  cosday2 |
| **Spatial factors** | | | Study area | Categorical, {Chikwawa/Ndirande/Chileka} (Reference: Chileka) | polygon |
| **Outcome variables** | | | ESBL *E. coli* | Binary, {1 = Sample colonised with ESBL *E.coli*, 0 = Sample not colonised with ESBL *E. coli*} | ecoli |
|  |  |  | ESBL *K. pneumoniae* | Binary, {1 = Sample colonised with ESBL *K. pneumoniae*, 0 = Sample not colonised with ESBL *K. pneumoniae*} | kpneu |
