## Supplementary Table 2 for "Risk factors, temporal dependence, and seasonality of human ESBL-producing *E. coli* and *K. pneumoniae* colonisation in Malawi: a longitudinal model-based approach"

|  | Log-odds | P-value | Odds ratio (95% CI) |
| --- | --- | --- | --- |
| Reactive to HIV testing (vs non-reactive) | -0.063 | 0.700 | 0.939 (0.682-1.293) |
| **Unknown HIV status (vs non-reactive)*** | **-0.168** | **0.074** | **0.846 (0.704-1.017)** |
| **Recent use of antibiotics** | **0.062** | **0.137** | **1.063 (0.981-1.153)** |
| **Age** | **0.092** | **0.026** | **1.097 (1.011-1.189)** |
| **Being male (vs female)** | **-0.175** | **0.039** | **0.840 (0.711-0.992)** |
| Number of people living in the household | 0.003 | 0.935 | 1.003 (0.924-1.089) |
| **Average household monthly income** | **-0.107** | **0.019** | **0.899 (0.822-0.983)** |
| Presence of a toilet in the household | -0.021 | 0.626 | 0.979 (0.900-1.066) |
| **Open defecation** | **0.089** | **0.038** | **1.093 (1.005-1.189)** |
| Sharing the toilet with non-household members | 0.017 | 0.699 | 1.017 (0.935-1.105) |
| **Presence of a disposal mechanism for animal waste** | **-0.090** | **0.046** | **0.914 (0.837-0.998)** |
| Eating street food | 0.054 | 0.214 | 1.055 (0.970-1.148) |
| **Eating from shared plates** | **-0.079** | **0.072** | **0.924 (0.848-1.007)** |
| **Having a pipe as drinking water source** | **-0.099** | **0.022** | **0.906 (0.832-0.985)** |
| Having a communal tap as drinking water source | -0.035 | 0.463 | 0.965 (0.879-1.061) |
| **Having a well as drinking water source** | **0.191** | **3.8e-04** | **1.210 (1.089-1.345)** |
| **Use of alternative water for cleaning utensils** | **0.057** | **0.174** | **1.059 (0.975-1.149)** |
| Owning birds | 4.1e-05 | 0.999 | 1.000 (0.915-1.094) |
| **Owning cattle, goats or sheep** | **0.093** | **0.044** | **1.097 (1.003-1.201)** |
| Owning dogs or cats | -0.020 | 0.628 | 0.980 (0.903-1.064) |
| Owning pigs | -0.038 | 0.397 | 0.962 (0.881-1.052) |
| **Keeping animals inside** | **0.122** | **0.005** | **1.129 (1.038-1.228)** |
| **Contact with river water** | **0.092** | **0.044** | **1.097 (1.003-1.200)** |
| Contact with drains | -0.034 | 0.430 | 0.967 (0.890-1.051) |
| Toilet type: other (vs no toilet) | -0.145 | 0.440 | 0.865 (0.600-1.249) |
| Toilet type: pit latrine (vs no toilet) | -0.026 | 0.826 | 0.974 (0.773-1.228) |
| Toilet type: shared toilet (vs no toilet) | -0.062 | 0.760 | 0.940 (0.630-1.402) |
| **Toilet floor material: no toilet (vs concrete/wood)** | **0.275** | **0.050** | **1.317 (1.000-1.736)** |
| **Toilet floor material: soil (vs concrete/wood)** | **0.330** | **0.002** | **1.391 (1.130-1.712)** |
| **Having a drop hole cover on the toilet** | **-0.164** | **1.7e-04** | **0.849 (0.779-0.925)** |
| Presence of toilet paper in the toilet | -0.053 | 0.234 | 0.948 (0.868-1.035) |
| **Presence of newspaper/paper in the toilet** | **-0.141** | **0.002** | **0.868 (0.796-0.947)** |
| Visible human faeces around the household | -0.009 | 0.830 | 0.991 (0.911-1.078) |
| Presence of handwashing facilities in the household | 0.037 | 0.405 | 1.038 (0.951-1.132) |
| **Frequency of soap presence in handwashing facilities** | **-0.082** | **0.079** | **0.921 (0.841-1.009)** |
| **Storing water covered** | **-0.067** | **0.098** | **0.935 (0.863-1.013)** |
| Storing water uncovered | 0.034 | 0.481 | 1.035 (0.941-1.137) |
| **Storing water in a container with lid/tap** | **-0.112** | **0.013** | **0.894 (0.819-0.976)** |
| **Contact between animals and food areas** | **0.187** | **1.3e-05** | **1.205 (1.108-1.311)** |
| Visible animal faeces around the household | -0.006 | 0.902 | 0.994 (0.904-1.093) |
| **Presence of standing water around the household** | **-0.069** | **0.121** | **0.933 (0.855-1.018)** |
| Having children of school age | -0.015 | 0.716 | 0.985 (0.908-1.069) |
| **Number of days since the first sample** | **0.118** | **0.005** | **1.125 (1.037-1.221)** |
| **Harmonic term (sinday)** | **-0.181** | **0.004** | **0.834 (0.738-0.943)** |
| **Harmonic term (cosday)** | **0.305** | **2e-04** | **1.357 (1.156-1.593)** |
| Harmonic term (sinday2) | -0.032 | 0.626 | 0.969 (0.852-1.102) |
| Harmonic term (cosday2) | -0.029 | 0.679 | 0.972 (0.848-1.113) |

*Variables under the threshold (0.2) highlighted in bold
