## Supplementary Table 4 for "Risk factors, temporal dependence, and seasonality of human ESBL-producing *E. coli* and *K. pneumoniae* colonisation in Malawi: a longitudinal model-based approach"

|  | Log-odds | P-value | Odds ratio (95% CI) |
| --- | --- | --- | --- |
| Reactive to HIV testing (vs non-reactive) | 0.104 | 0.661 | 1.109 (0.697-1.765) |
| Unknown HIV status (vs non-reactive) | 0.041 | 0.750 | 1.042 (0.809-1.342) |
| **Recent use of antibiotics*** | **0.107** | **0.058** | **1.112 (0.996-1.242)** |
| Age | -0.019 | 0.758 | 0.981 (0.868-1.108) |
| Being male (vs female) | -0.159 | 0.207 | 0.853 (0.666-1.092) |
| **Number of people living in the household** | **0.229** | **6.1e-05** | **1.257 (1.124-1.406)** |
| Average household monthly income | -0.014 | 0.830 | 0.987 (0.872-1.116) |
| **Presence of a toilet in the household** | **0.085** | **0.193** | **1.088 (0.958-1.236)** |
| Open defecation | 0.019 | 0.755 | 1.019 (0.904-1.150) |
| Sharing the toilet with non-household members | 0.055 | 0.371 | 1.056 (0.937-1.191) |
| Presence of a disposal mechanism for animal waste | 0.057 | 0.330 | 1.058 (0.944-1.186) |
| **Eating street food** | **-0.125** | **0.029** | **0.882 (0.788-0.987)** |
| **Eating from shared plates** | **-0.152** | **0.016** | **0.859 (0.759-0.972)** |
| **Having a pipe as drinking water source** | **0.106** | **0.077** | **1.112 (0.989-1.250)** |
| **Having a communal tap as drinking water source** | **-0.120** | **0.072** | **0.887 (0.778-1.011)** |
| Having a tube/well as drinking water source | -0.009 | 0.891 | 0.992 (0.878-1.119) |
| **Use of alternative water for cleaning utensils** | **-0.087** | **0.187** | **0.917 (0.806-1.043)** |
| **Owning birds** | **0.138** | **0.026** | **1.148 (1.016-1.296)** |
| Owning cattle, goats or sheep | 0.054 | 0.370 | 1.056 (0.938-1.188) |
| **Owning dogs or cats** | **0.086** | **0.153** | **1.089 (0.969-1.225)** |
| **Owning pigs** | **0.085** | **0.131** | **1.089 (0.975-1.216)** |
| Keeping animals inside | 0.041 | 0.506 | 1.042 (0.924-1.174) |
| Contact with river water | -0.001 | 0.986 | 0.999 (0.885-1.128) |
| **Contact with drains** | **0.137** | **0.011** | **1.147 (1.032-1.275)** |
| Toilet type: other (vs no toilet) | 0.276 | 0.299 | 1.317 (0.783-2.216) |
| **Toilet type: pit latrine** **(vs no toilet)** | **0.287** | **0.107** | **1.333 (0.940-1.889)** |
| Toilet type: shared toilet (vs no toilet) | -0.396 | 0.261 | 0.673 (0.338-1.343) |
| Toilet floor material: no toilet (vs concrete/wood) | -0.148 | 0.454 | 0.863 (0.586-1.270) |
| Toilet floor material: soil (vs concrete/wood) | 0.154 | 0.270 | 1.167 (0.887-1.534) |
| Having a drop hole cover on the toilet | 0.042 | 0.488 | 1.043 (0.926-1.176) |
| Presence of toilet paper in the toilet | 0.025 | 0.676 | 1.026 (0.910-1.156) |
| Presence of newspaper/paper in the toilet | -0.047 | 0.459 | 0.954 (0.842-1.081) |
| **Visible human faeces around the household** | **0.109** | **0.074** | **1.115 (0.990-1.256)** |
| Presence of handwashing facilities in the household | 0.060 | 0.341 | 1.062 (0.939-1.201) |
| Frequency of soap presence in handwashing facilities | -0.064 | 0.344 | 0.938 (0.822-1.071) |
| Storing water covered | 0.080 | 0.280 | 1.083 (0.937-1.252) |
| **Storing water uncovered** | **-0.102** | **0.093** | **0.903 (0.801-1.017)** |
| Storing water in a container with lid/tap | -0.007 | 0.910 | 0.993 (0.879-1.121) |
| Contact between animals and food areas | -0.050 | 0.429 | 0.952 (0.842-1.076) |
| Visible animal faeces around the household | -0.025 | 0.678 | 0.975 (0.865-1.099) |
| Presence of standing water around the household | -0.047 | 0.463 | 0.954 (0.843-1.081) |
| Having children of school age | 0.009 | 0.882 | 1.009 (0.893-1.140) |
| **Number of days since the first sample** | **-0.097** | **0.122** | **0.907 (0.802-1.026)** |
| **Harmonic term (sinday)** | **-0.345** | **3.7e-04** | **0.708 (0.586-0.856)** |
| **Harmonic term (cosday)** | **0.243** | **0.038** | **1.275 (1.014-1.605)** |
| Harmonic term (sinday2) | -0.053 | 0.592 | 0.948 (0.781-1.152) |
| **Harmonic term (cosday2)** | **0.166** | **0.095** | **1.181 (0.972-1.434)** |
| **Living in Chikwawa (vs Chileka)** | **0.196** | **0.183** | **1.216 (0.912-1.622)** |
| Living in Ndirande (vs Chileka) | 0.098 | 0.536 | 1.103 (0.809-1.504) |

*Variables under the threshold (0.2) highlighted in bold
