## Supplementary figures and images for "Risk factors, temporal dependence, and seasonality of human ESBL-producing *E. coli* and *K. pneumoniae* colonisation in Malawi: a longitudinal model-based approach"

### Supplementary Figure 1

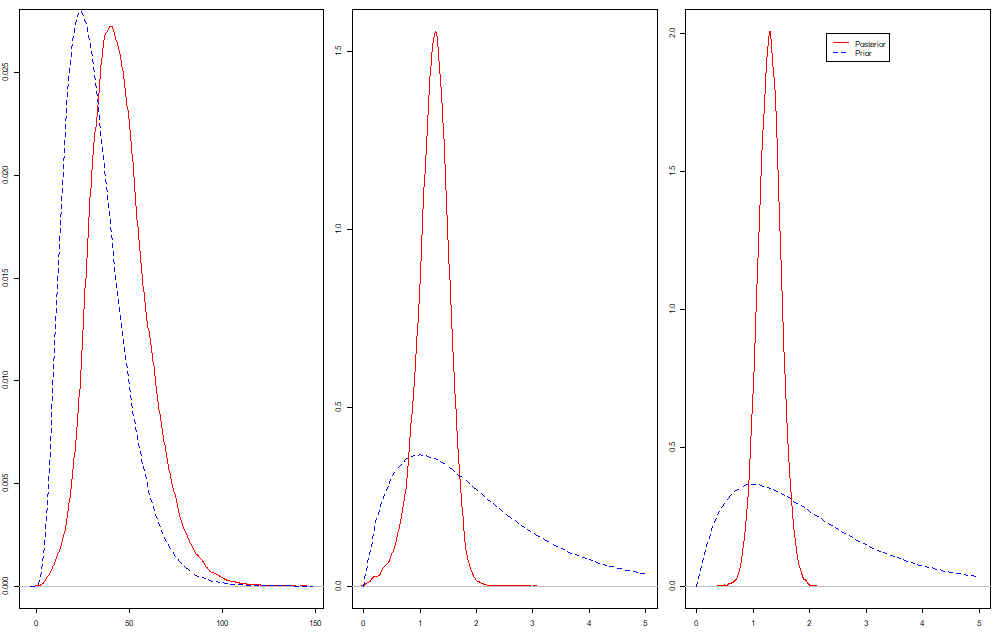

### Supplementary Figure 2

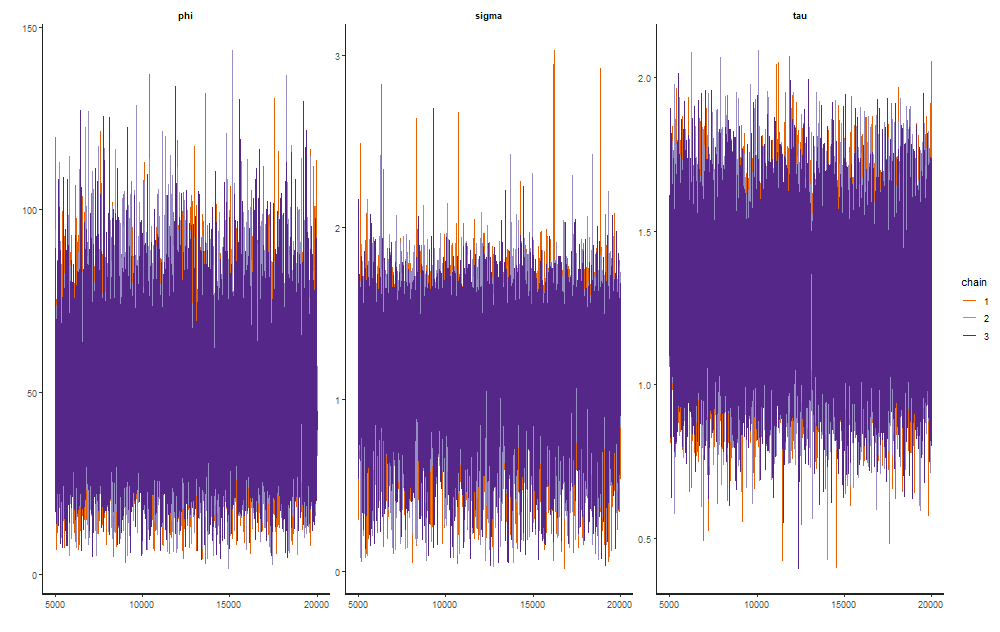

### Supplementary Figure 3

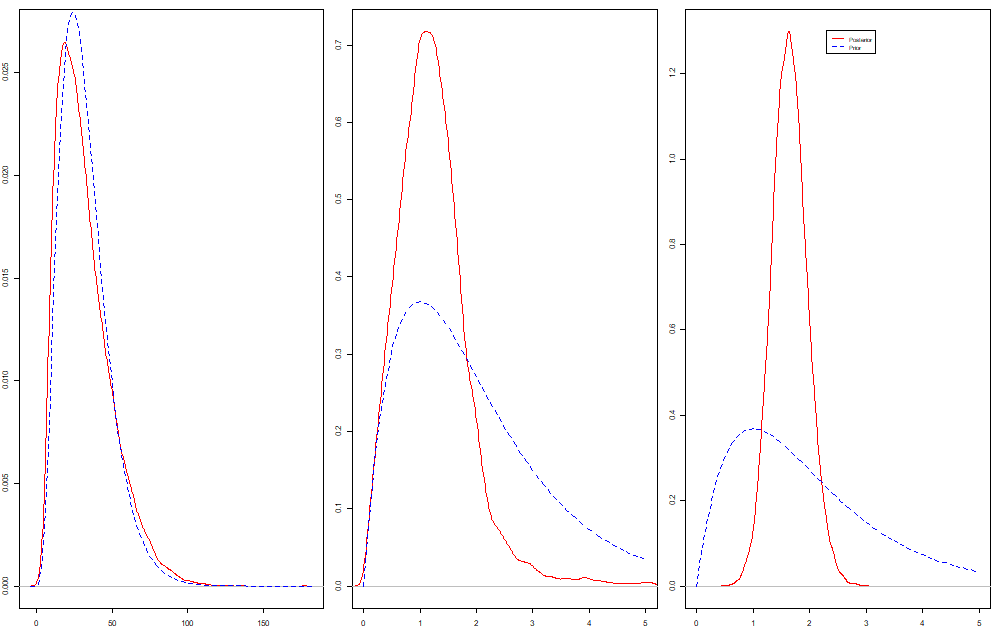

### Supplementary Figure 4

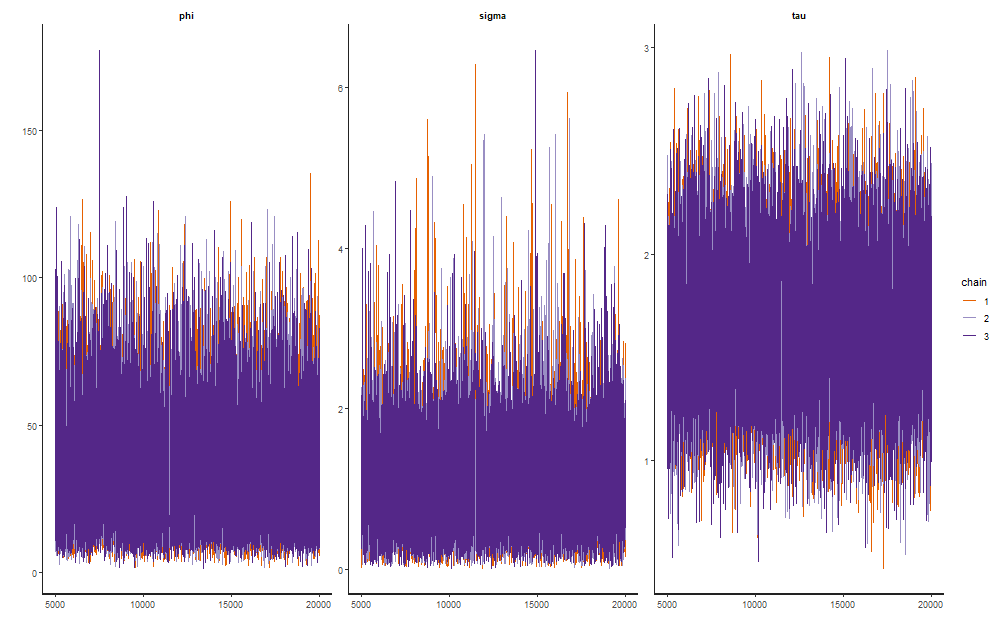
